# Analysis of hospitalized COVID-19 patients in the Mount Sinai Health System using electronic medical records (EMR) reveals important prognostic factors for improved clinical outcomes

**DOI:** 10.1101/2020.04.28.20075788

**Authors:** Zichen Wang, Amanda B. Zheutlin, Yu-Han Kao, Kristin L. Ayers, Susan J. Gross, Patricia Kovatch, Sharon Nirenberg, Alexander W. Charney, Girish N. Nadkarni, Paul F. O’Reilly, Allan C. Just, Carol R. Horowitz, Glenn Martin, Andrea D. Branch, Benjamin S. Glicksberg, Dennis S. Charney, David L. Reich, William K. Oh, Eric E. Schadt, Rong Chen, Li Li

## Abstract

**Importance:** There is an urgent need to understand patient characteristics of having COVID-19 disease and evaluate markers of critical illness and mortality.

**Objective:** To assess association of clinical features on patient outcomes.

**Design, Setting, and Participants:** In this observational case series, patient-level data were extracted from electronic medical records for 28,336 patients tested for SARS-CoV-2 at the Mount Sinai Health System from 2/24/ to 4/15/2020, including 6,158 laboratory-confirmed cases.

**Exposures:** Confirmed COVID-19 diagnosis by RT-PCR assay from nasal swabs.

**Main Outcomes and Measures:** Effects of race on positive test rates and mortality were assessed. Among positive cases admitted to the hospital (N = 3,273), effects of patient demographics, hospital site and unit, social behavior, vital signs, lab results, and disease comorbidities on discharge and death were estimated.

**Results:** Hispanics (29%) and African Americans (25%) had disproportionately high positive case rates relative to population base rates (*p*<2e-16); however, no differences in mortality rates were observed in the hospital. Outcome differed significantly between hospitals (Gray’s T=248.9; *p*<2e-16), reflecting differences in average baseline age and underlying comorbidities. Significant risk factors for mortality included age (HR=1.05 [95% CI, 1.04-1.06]; p=1.15e-32), oxygen saturation (HR=0.985 [95% CI, 0.982-0.988]; p=1.57e-17), care in ICU areas (HR=1.58 [95% CI, 1.29-1.92]; p=7.81e-6), and elevated creatinine (HR=1.75 [95% CI, 1.47-2.10]; p=7.48e-10), alanine aminotransferase (ALT) (HR=1.002, [95% CI 1.001-1.003]; p=8.86e-5) white blood cell (WBC) (HR=1.02, [95% CI 1.01-1.04]; p=8.4e-3) and body-mass index (BMI) (HR=1.02, [95% CI 1.00-1.03]; p=1.09e-2). Asthma (HR=0.78 [95% CI, 0.62-0.98]; p=0.031) was significantly associated with increased length of hospital stay, but not mortality. Deceased patients were more likely to have elevated markers of inflammation. Baseline age, BMI, oxygen saturation, respiratory rate, WBC count, creatinine, and ALT were significant prognostic indicators of mortality.

**Conclusions and Relevance:** While race was associated with higher risk of infection, we did not find a racial disparity in inpatient mortality suggesting that outcomes in a single tertiary care health system are comparable across races. We identified clinical features associated with reduced mortality and discharge. These findings could help to identify which COVID-19 patients are at greatest risk and evaluate the impact on survival.

## Introduction

Coronavirus disease 2019 (COVID-19) is a global pandemic that has already infected over 2 million individuals, including over 630,000 in the US as of Apr 15, 2020^1^. Given recent reports on the high rate of COVID-19 infections among those who remain asymptomatic, however, the true rate of infection is expected to be significantly higher than reported^1,2^. More than 26,000 US residents have died, the majority in epicenters like New York City (NYC), where 118,302 cases and 8,455 deaths have occurred to date^1^. Mortality rates have been disproportionately high among Hispanic and African American individuals. In NYC, death rates for these groups are nearly double those in Caucasians or Asians, but the factors contributing to this racial disparity remain unclear^2^. Furthermore, reducing mortality among all critically ill individuals is the highest priority, although some clinical risk factors have been noted in recent publications^3,4^, there remains much to learn about which patients are at highest risk, what factors are most indicative of disease progression and prognosis, and which interventions may be effective.

A systematic review of studies predicting coronavirus-related outcomes concluded that all of the publications were biased in some way, limiting their utility in practice^3^. They noted that prognostic models often excluded patients for which no outcome was yet determined (e.g., patients that had neither recovered nor died) leading to selection bias, used relatively small samples (e.g., 26–577 patients) increasing risk of overfitting, or in many cases did not use features or timepoints that could be measured prospectively (e.g., last available vital sign)^3^. Furthermore, none of these studies were conducted in the US where population factors, health-related behaviors, cultural differences, and hospital standard of care protocols may be different^4^. Indeed, case characteristics among hospitalized patients in China seem meaningfully different than those in the US^5^ – for example, mortality and certain mortality rates were much lower.

There have been several recent descriptive reports of COVID-19 clinical characteristics among patients admitted to US hospitals^6^, including an ongoing, population-based report from the Center for Disease Control and Prevention^7^ and another from NYU Langone Health^5^. While data is mounting regarding racial disparities and COVID-19^5^ most of these studies have not offered in depth analysis by race to investigate racial disparities in mortality. Since no proven effective therapies exist for COVID-19^6^, up-to-date clinical outcomes and data on factors influencing risk for mortality and recovery over time are urgently needed.

Given the high mortality rate in NYC and the uncertainty of COVID-19 progression during inpatient stays, accurately evaluating prognosis of both mortality and discharge among hospitalized individuals is critically needed. Identifying which patients are at highest risk for mortality will enable clinicians to target interventions, allocate resources, and make more informed triage decisions. Highlighting factors most associated with mortality will also help prioritize factors to monitor during hospital admission. Prognostic models have the potential to improve the standard of care for COVID-19, open opportunities for testing investigational drugs in clinical trials, and guide clinical acumen.

In this study, the largest and most racially diverse US-based COVID-19 case series to date, we provided descriptive statistics on laboratory-confirmed cases (N = 6,158) and hospitalized patients (N = 3,273) from the Mount Sinai Health System (MSHS). In particular, we investigated the impact of race on mortality given the racial disparities in mortality rates observed in the US^2^. We also evaluated the effects of demographics, social behavior, and clinical variables on mortality and recovery among hospitalized patients admitted to one of MSHS hospitals in Manhattan, Brooklyn, and Queens. As in previous studies^5–7^, we considered disease comorbidities, vital signs, and lab results for COVID-19 disease and potential to improve patient outcomes. We reported hazard ratios for all risk factors estimated using a competing risk model that simultaneously considered two outcomes resulting in hospitalization termination, death and discharge. We also developed a prognostic model using exclusively data available at baseline, the first such model in a diverse NYC with diverse population. Together, we believe these estimates can help inform stakeholders which COVID-19 patients are at greatest risk for poor outcomes and evaluate the impact on survival.

## Methods

### Electronic Medical Record data and processing

This study utilized the de-identified EMR data and was exempted from the Mount Sinai Institutional Review Board due to the COVID-19 pandemic. We obtained de-identified data from the electronic medical record (EMR; EPIC Systems) via the Mount Sinai Data Warehouse through April 15, 2020. The EMR dataset included patients with at least one encounter at a Mount Sinai facility who had been diagnosed with COVID-19, who were under investigation for COVID-19 or who had been screened negative for COVID-19. Patients included in the dataset were those who had an encounter (either in person or virtual) at a Mount Sinai facility in which a COVID-19 test was ordered or a COVID-19 related diagnosis, including suspected diagnosis like ‘Suspected 2019-Ncov diagnosis’, was given.

In total, 28,336 patients were included in the de-identified EMR dataset. Demographics including age, sex, race, ethnicity and smoking status, as well as disease comorbidities were extracted from the EMR. Comorbidities were defined as the presence or absence of the following chronic conditions recorded as “Active” in the Epic Problem List: diabetes mellitus, hypertension, asthma, chronic obstructive pulmonary disease (COPD), human immunodeficiency virus (HIV) infection, obesity and cancer. For each encounter, initial measurements of vital signs including BMI, temperature, systolic blood pressure (BP), diastolic BP, O_2_ saturation, heart rate and respiratory rate were provided. Laboratory test orders and results throughout these encounters were also extracted; common lab test orders included complete blood count (CBC) and differentials, metabolic panels, blood lactate dehydrogenase (LDH), ferritin, fibrin degradation dimer (D-Dimer), serum procalcitonin, hepatic function panel, blood culture, fibrinogen, C reactive protein (CRP).

Since vital signs can shift during a single encounter, we also extracted the maximum temperature and minimum O_2_ saturation for each encounter, as well as the length of stay for inpatients.

### COVID-19 case definition

A confirmed case of COVID-19 was defined as a positive test result from real-time reverse-transcriptase-polymerase-chain-reaction (RT-PCR) assay nasopharyngeal swab specimens. A patient under investigation (PUI) of COVID-19 was clinically defined as patients who experienced 1) fever and/or cough, shortness-of-breath (SOB), sore throat, nasal congestion not related to typical seasonal allergies or 2) fever and/or cough and a history of exposure to COVID-19. Collectively, our study included 6,158 COVID-19 patients and 428 PUIs with confirmed positive or presumptive positive COVID-19 RT-PCR test results.

### Examination of racial disparity in diagnosis and mortality rates

We compiled a background population from the MSHS EMR as a demographic reference for the residents in the areas of NYC served by Mount Sinai. We selected all patients with encounters of any types (outpatient, inpatient, emergency) recorded in the MSHS EMR since 2016 and retrieved their self-reported races, ethnicities and age in years. Patients with unknown race/ethnicity information were excluded from this analysis. In total, the background population was composed of 1.6 million people, with 48.1% white, 21.6% African American/Black, 15.3% other, 8.5% Asian/Pacific Islander and 6.4% Hispanic/Latinos (Table 1).

**Table 1.** Constituents of patients in the background MSHS EMR, patients tested for SARS CoV-2 infection, SARS-CoV-2 infected patients and patients deceased from Covid-19.

| RACE | BACKGROUND | TESTED | INFECTED | DECEASED |
| --- | --- | --- | --- | --- |
| AFRICAN-AMERICAN/BLACK | 360542 (21.6%) | 4744 (19.4%) | 1528 (24.6%) | 235 (24.1%) |
| ASIAN/PACIFIC-ISLANDER | 142080 (8.5%) | 1679 (6.9%) | 318 (5.1%) | 43 (4.4%) |
| CAUCASIAN/WHITE | 802848 (48.1%) | 9194 (37.6%) | 1642 (26.5%) | 293 (30.1%) |
| HISPANIC/LATINO | 107255 (6.4%) | 5722 (23.4%) | 1769 (28.5%) | 244 (25.1%) |
| OTHER | 254682 (15.3%) | 3100 (12.7%) | 947 (15.3%) | 159 (16.3%) |

Next, we tallied the numbers of SARS-CoV-2 infected patients defined previously and those who were deceased as of April 15, 2020 (Table 1). To test for racial disparities in positive test frequencies and COVID-19 mortality, we compared observed rates of COVID-19 diagnosis and mortality relative to the MSHS race frequencies. We performed a Chi-square test of independence of variables in contingency tables under the null hypothesis that the COVID-19 diagnosis rates or mortality rates were not different among the five racial groups. We also fitted a multivariate logistic regression (implemented in ‘glm’ function in R version 3.6.1) to adjust for confounding variables such as age on diagnosis rates. Finally, competing risks survival analysis was employed to analyze the mortality rate and discharge rate over time.

### Hospitalized COVID-19 patient cohort

We included COVID-19 patients (N=3,061) and PUIs (N=212) with confirmed positive or presumptive positive COVID-19 RT-PCR test results who were admitted as inpatients and stayed at least one day in the hospital. We recorded the durations of hospital stay and the number of days since SARS-CoV-2 positive. The diagnosis day was defined as the date when the COVID-19 test with positive results was ordered. Three possible outcomes were defined for our hospitalized COVID-19 cohort: in-hospital death (deceased), discharged to home or other locations not concerning intensive medical care (recovered), and continued hospitalization (right-censored). For individual patients, we extracted all hospital encounters up to April 15, 2020 that were related to COVID-19 diagnostic tests or hospital admissions.

### Analyses of individual factors on inpatient mortality over time

We were first interested in observing any demographic or hospital site- or specialty-specific effects on mortality and discharge individually over time. To do this, we estimated the cumulative incidence functions (CIFs) for in-hospital death and discharge using a univariate competing risks survival analysis for each covariate individually: race, sex, hospital, and care area within the hospital.

### Multivariate regression among inpatients with patient outcomes

Next, we assessed which factors were associated with death among hospitalized patients. For this analysis, we only included subjects with known outcomes (death: N = 742; discharge: N = 1,706). We fitted a multivariate logistic regression on clinical outcome (deceased=1, recovered =0) using duration of stay, demographic factors, vital signs, comorbidities, care in ICU unit, and common laboratory tests ordered at time of hospital admission.

### Multivariate analyses on inpatient mortality: Competing risk survival analysis

To assess the impact of clinical variables on survival, we modeled the outcomes of hospitalized COVID-19 patients using competing risks survival analyses, which treats the two distinctive outcomes, in-hospital death and cured as two competing causes for the same event, i.e. termination of hospitalization. Competing risks models are recommended over Kaplan-Meier survival analysis for studying events with multiple underlying causes^7–9^. More formally, we denote the duration of hospital stay since diagnosis as *t* and the total duration from diagnosis to termination of hospitalization as *t_f_*. The goal of competing risks survival analysis is to estimate the CIF for each individual cause k. CIF is a function of time defined as the probability of a patient who stayed at hospital for a duration of *t_f_*:

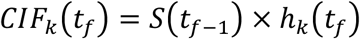

, where *S*(*t*) is the survival function and ℎ*_k_*(*t*) is the hazard function for the cause *k*. To estimate the CIFs for in-hospital death and recovery for COVID-19 patients, we used the “cuminc” function in the R package “cmprsk”. Gray’s test^10^ was conducted to determine if there were statistical differences among CIFs corresponding to subgroups of patients, under the null hypothesis that the CIFs under consideration were not different from each other.

To identify covariates associated with the two clinical outcomes – represented in the covariate vector ***x*** – we performed multivariate statistical analyses to estimate the contribution of each potential covariate to the cause-specific hazard function ℎ*_k_*(*t*) as follows:

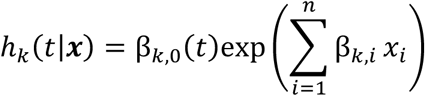

, where β*_k_*_,0_(*t*) is the baseline hazard of case *k*, and β*_k,i_* is the coefficient for covariate *i*. When estimating the β*_k,i_* for competing risks analyses, we applied two families of models: cause-specific hazard models and subdistribution hazard models^11^. Cause-specific hazard models were estimated using the Cox proportional hazard approach^12^ using the “coxph” function in the R package “survival” by treating one cause as the event and the other as right-censored, whereas subdistribution hazard models were fitted using the “cmr” function in the R package “cmprsk”^13^. The R software used throughout this research is in version 3.6.1.

### Prognostic forecast of inpatient mortality

To help with prediction, we performed the same analysis used for estimating effects on survival described already (“Multivariate analyses on inpatient mortality: Competing risk survival analysis”), but limited features to only those available at baseline when patients were admitted to the hospital.

## Results

Our cohort included 28,336 individuals tested (Table 1) for COVID-19. 6,158 (21.7%) patients tested positive and an additional 428 (1.5%) were presumed positive and thus included as cases by our definition. 3,273 cases out of all positives (53.2%) were admitted to one of five hospitals included in the Mount Sinai Health System which use the EPIC EMR system. Of those, 742 died (22.6%), 1,706 were discharged (52%), and 825 remained hospitalized (25.2%) at the end point of this study (April 15, 2020).

We investigated the impact of race and age on COVID-19 diagnosis rates and mortality. We then assessed the influence of a variety of factors on inpatient mortality. First, we estimated the individual effect of race, sex, hospital, and specialty unit on inpatient mortality over time. Next, we identified the strongest predictors of inpatient mortality considering all possible indicators regardless of time, which can help prioritize clinical features to monitor. Then, to assess the impact of each feature on survival, we estimated hazard ratios from a competing risks survival analysis. Finally, to offer a prognostic tool for prioritizing highest risk patients, we used baseline information to forecast mortality.

### Racial disparity for SARS-CoV-2 positive: Hispanics and African Americans have elevated COVID-19 diagnosis rates

We found Hispanics and African Americans were over-represented in the SARS-CoV-2 infected cohort, accounting for 28.5% and 24.6%, respectively, of all the infected patients with known self-reported races, which were both significantly higher rates than expected based on the population base rate (Chi2=5340.5; *p* < 2e-16; Table 1). We also noted the age distributions of infected patients within each race were different relative to our reference population (Fig. S1A). Furthermore, the Caucasian deceased cohort shows a different age distribution from the other race groups. Its density increases continuously as age increases, forming a triangle shape, while African Americans shows the highest density at the age of 74 years (Fig S1B). However, even after adjusting for age in logistic regression, we found that Hispanics, African Americans and people identified as other races have significantly higher odds of being infected by SARS-CoV-2 compared to Caucasian individuals (Table S1). Age-adjusted COVID-19 diagnosis rates in Caucasian and Asian American groups were not statistically different. Together, this has clearly demonstrated that some minority groups, including Hispanics and African Americans, were at higher risk of being infected by SARS-CoV-2 in the New York metropolitan area served by the MSHS.

Interestingly, we found a bimodal age distribution in infected cohorts of each race, with the first peak in the age group of 20–40 years and second peak in the age group of 50–70 (Fig S1A). Within the deceased population, Caucasians showed a triangular shape of age distribution with higher density when age increases, while African Americans showed the highest density around the age of 75 years (Fig S1B).

### No racial disparity is detected among COVID-19 patients for in-hospital mortality

Importantly we found no differences in mortality rates across all racial groups (*p* =0.056; Table 1). We also found that older age increased risk for mortality (Fig. 1), as has been previously reported elsewhere^3,4,6^. Furthermore, we did not find an effect of race on clinical outcome after adjusting for underlying covariates (Fig. 3), which further suggests racial parity in terms of inhospital mortality in our cohort.

**Figure 1.**
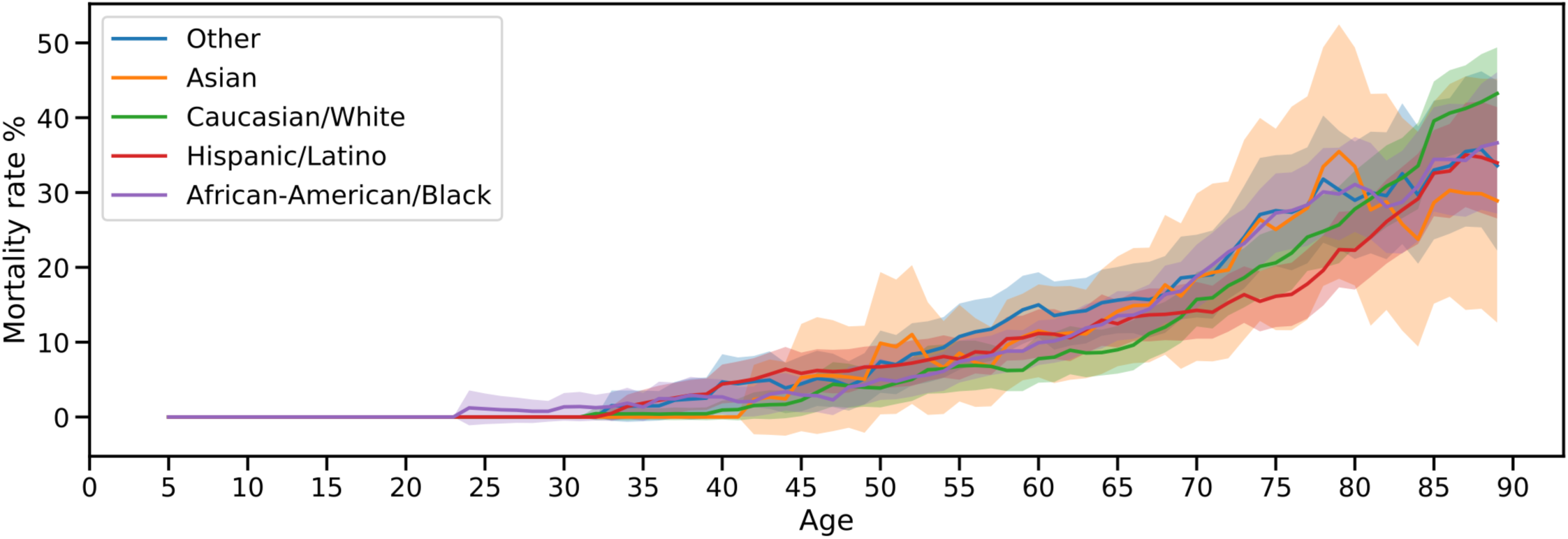
In-hospital mortality rates of COVID-19 patients break-down by self-reported races and age groups. In-hospital mortality rate is defined by the number of deceased patients during hospitalization divided by the total number of Covid-19 patients and PUIs in our cohort. The mortality rates of different age groups are plotted across different racial groups indicated in the legend. The 95% confidence intervals (CI) for the mortality rates across race groups are estimated using bootstrap by sampling the patients in that a rolling 10-year age window 500 times.

### Factors influencing inpatient mortality

To identify factors involved in the progression and prognosis of the disease, we focused on COVID-19 patients admitted to the hospital. Our hospitalized COVID-19 cohort contains 3,273 patients with at least one day in the hospital, among whom 742 died, 1,706 were discharged (presumed recovered), and 825 were still hospitalized as of April 15, 2020 (Table 2). We summarize the demographic features, comorbidities, vital signs and laboratory tests at admission, as well as the distribution of patients by hospital site, care in the intensive care unit (ICU) area, and number of days from diagnosis to discharge or last follow-up (Table 2).

**Table 2.**
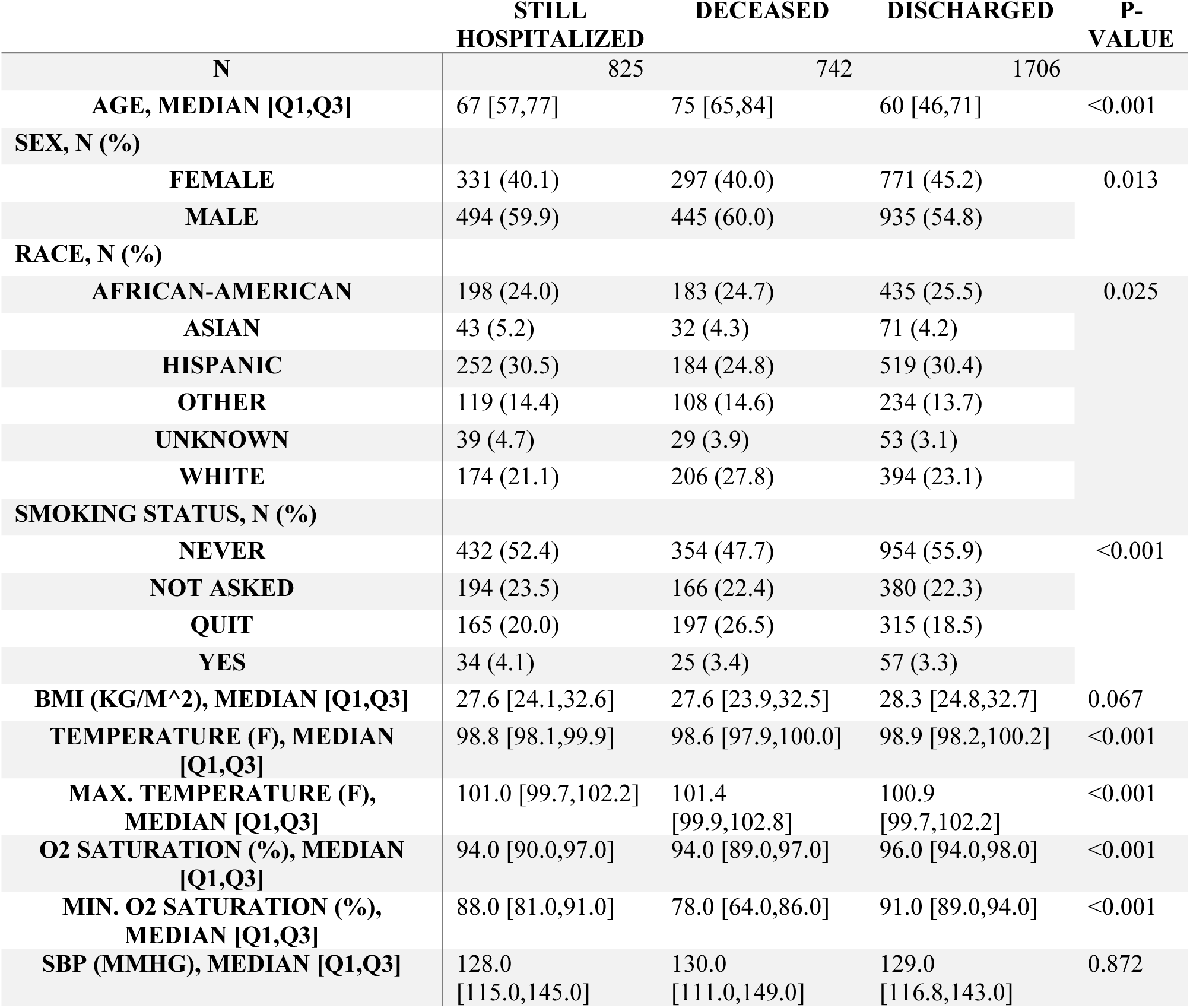
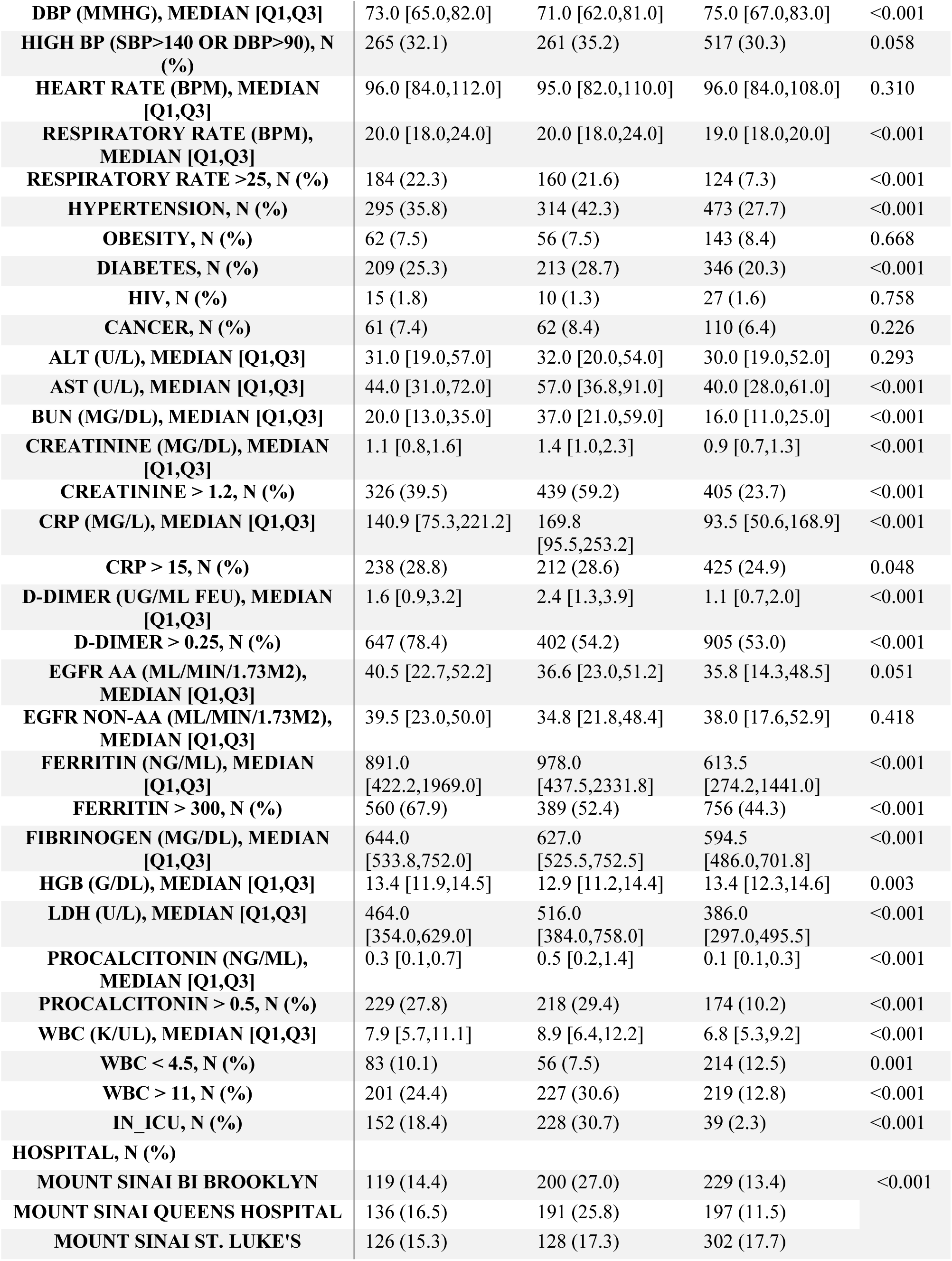
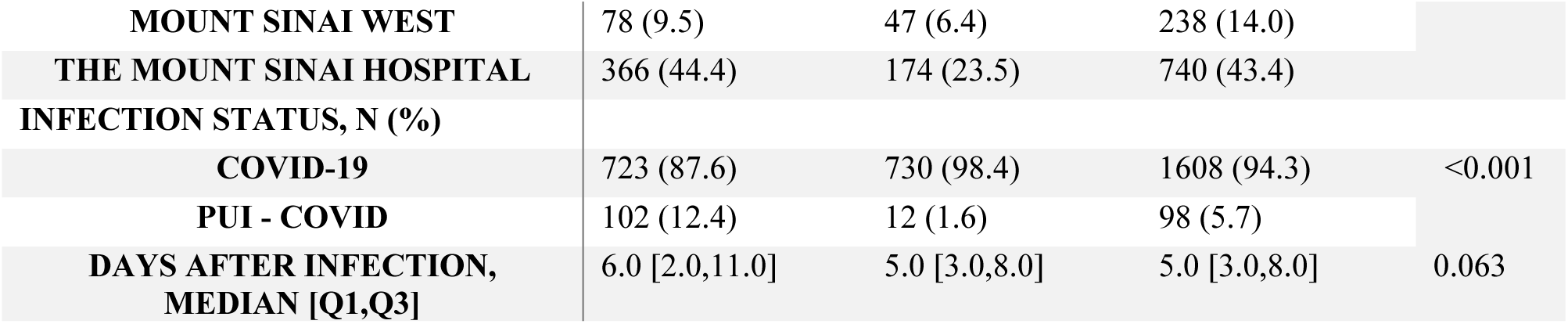
Characteristics of the hospitalized Covid-19 cohort grouped by their outcomes.

|  | STILL<br>HOSPITALIZED | DECEASED | DISCHARGED | P-VALUE |
| --- | --- | --- | --- | --- |
| N | 825 | 742 | 1706 |  |
| AGE, MEDIAN [Q1,Q3] | 67 [57,77] | 75 [65,84] | 60 [46,71] | <0.001 |
| SEX, N (%) |  |  |  |  |
| FEMALE | 331 (40.1) | 297 (40.0) | 771 (45.2) | 0.013 |
| MALE | 494 (59.9) | 445 (60.0) | 935 (54.8) |  |
| RACE, N (%) |  |  |  |  |
| AFRICAN-AMERICAN | 198 (24.0) | 183 (24.7) | 435 (25.5) | 0.025 |
| ASIAN | 43 (5.2) | 32 (4.3) | 71 (4.2) |  |
| HISPANIC | 252 (30.5) | 184 (24.8) | 519 (30.4) |  |
| OTHER | 119 (14.4) | 108 (14.6) | 234 (13.7) |  |
| UNKNOWN | 39 (4.7) | 29 (3.9) | 53 (3.1) |  |
| WHITE | 174 (21.1) | 206 (27.8) | 394 (23.1) |  |
| SMOKING STATUS, N (%) |  |  |  |  |
| NEVER | 432 (52.4) | 354 (47.7) | 954 (55.9) | <0.001 |
| NOT ASKED | 194 (23.5) | 166 (22.4) | 380 (22.3) |  |
| QUIT | 165 (20.0) | 197 (26.5) | 315 (18.5) |  |
| YES | 34 (4.1) | 25 (3.4) | 57 (3.3) |  |
| BMI (KG/M^2), MEDIAN [Q1,Q3] | 27.6 [24.1,32.6] | 27.6 [23.9,32.5] | 28.3 [24.8,32.7] | 0.067 |
| TEMPERATURE (F), MEDIAN [Q1,Q3] | 98.8 [98.1,99.9] | 98.6 [97.9,100.0] | 98.9 [98.2,100.2] | <0.001 |
| MAX. TEMPERATURE (F), MEDIAN [Q1,Q3] | 101.0 [99.7,102.2] | 101.4 [99.9,102.8] | 100.9 [99.7,102.2] | <0.001 |
| O2 SATURATION (%), MEDIAN [Q1,Q3] | 94.0 [90.0,97.0] | 94.0 [89.0,97.0] | 96.0 [94.0,98.0] | <0.001 |
| MIN. O2 SATURATION (%), MEDIAN [Q1,Q3] | 88.0 [81.0,91.0] | 78.0 [64.0,86.0] | 91.0 [89.0,94.0] | <0.001 |
| SBP (MMHG), MEDIAN [Q1,Q3] | 128.0 [115.0,145.0] | 130.0 [111.0,149.0] | 129.0 [116.8,143.0] | 0.872 |
| **DBP (MMHG), MEDIAN [Q1,Q3]** | 73.0 [65.0,82.0] | 71.0 [62.0,81.0] | 75.0 [67.0,83.0] | <0.001 |
| **HIGH BP (SBP>140 OR DBP>90), N (%)** | 265 (32.1) | 261 (35.2) | 517 (30.3) | 0.058 |
| **HEART RATE (BPM), MEDIAN [Q1,Q3]** | 96.0 [84.0,112.0] | 95.0 [82.0,110.0] | 96.0 [84.0,108.0] | 0.310 |
| **RESPIRATORY RATE (BPM), MEDIAN [Q1,Q3]** | 20.0 [18.0,24.0] | 20.0 [18.0,24.0] | 19.0 [18.0,20.0] | <0.001 |
| **RESPIRATORY RATE >25, N (%)** | 184 (22.3) | 160 (21.6) | 124 (7.3) | <0.001 |
| **HYPERTENSION, N (%)** | 295 (35.8) | 314 (42.3) | 473 (27.7) | <0.001 |
| **OBESITY, N (%)** | 62 (7.5) | 56 (7.5) | 143 (8.4) | 0.668 |
| **DIABETES, N (%)** | 209 (25.3) | 213 (28.7) | 346 (20.3) | <0.001 |
| **HIV, N (%)** | 15 (1.8) | 10 (1.3) | 27 (1.6) | 0.758 |
| **CANCER, N (%)** | 61 (7.4) | 62 (8.4) | 110 (6.4) | 0.226 |
| **ALT (U/L), MEDIAN [Q1,Q3]** | 31.0 [19.0,57.0] | 32.0 [20.0,54.0] | 30.0 [19.0,52.0] | 0.293 |
| **AST (U/L), MEDIAN [Q1,Q3]** | 44.0 [31.0,72.0] | 57.0 [36.8,91.0] | 40.0 [28.0,61.0] | <0.001 |
| **BUN (MG/DL), MEDIAN [Q1,Q3]** | 20.0 [13.0,35.0] | 37.0 [21.0,59.0] | 16.0 [11.0,25.0] | <0.001 |
| **CREATININE (MG/DL), MEDIAN [Q1,Q3]** | 1.1 [0.8,1.6] | 1.4 [1.0,2.3] | 0.9 [0.7,1.3] | <0.001 |
| **CREATININE > 1.2, N (%)** | 326 (39.5) | 439 (59.2) | 405 (23.7) | <0.001 |
| **CRP (MG/L), MEDIAN [Q1,Q3]** | 140.9 [75.3,221.2] | 169.8 [95.5,253.2] | 93.5 [50.6,168.9] | <0.001 |
| **CRP > 15, N (%)** | 238 (28.8) | 212 (28.6) | 425 (24.9) | 0.048 |
| **D-DIMER (UG/ML FEU), MEDIAN [Q1,Q3]** | 1.6 [0.9,3.2] | 2.4 [1.3,3.9] | 1.1 [0.7,2.0] | <0.001 |
| **D-DIMER > 0.25, N (%)** | 647 (78.4) | 402 (54.2) | 905 (53.0) | <0.001 |
| **EGFR AA (ML/MIN/1.73M2), MEDIAN [Q1,Q3]** | 40.5 [22.7,52.2] | 36.6 [23.0,51.2] | 35.8 [14.3,48.5] | 0.051 |
| **EGFR NON-AA (ML/MIN/1.73M2), MEDIAN [Q1,Q3]** | 39.5 [23.0,50.0] | 34.8 [21.8,48.4] | 38.0 [17.6,52.9] | 0.418 |
| **FERRITIN (NG/ML), MEDIAN [Q1,Q3]** | 891.0 [422.2,1969.0] | 978.0 [437.5,2331.8] | 613.5 [274.2,1441.0] | <0.001 |
| **FERRITIN > 300, N (%)** | 560 (67.9) | 389 (52.4) | 756 (44.3) | <0.001 |
| **FIBRINOGEN (MG/DL), MEDIAN [Q1,Q3]** | 644.0 [533.8,752.0] | 627.0 [525.5,752.5] | 594.5 [486.0,701.8] | <0.001 |
| **HGB (G/DL), MEDIAN [Q1,Q3]** | 13.4 [11.9,14.5] | 12.9 [11.2,14.4] | 13.4 [12.3,14.6] | 0.003 |
| **LDH (U/L), MEDIAN [Q1,Q3]** | 464.0 [354.0,629.0] | 516.0 [384.0,758.0] | 386.0 [297.0,495.5] | <0.001 |
| **PROCALCITONIN (NG/ML), MEDIAN [Q1,Q3]** | 0.3 [0.1,0.7] | 0.5 [0.2,1.4] | 0.1 [0.1,0.3] | <0.001 |
| **PROCALCITONIN > 0.5, N (%)** | 229 (27.8) | 218 (29.4) | 174 (10.2) | <0.001 |
| **WBC (K/UL), MEDIAN [Q1,Q3]** | 7.9 [5.7,11.1] | 8.9 [6.4,12.2] | 6.8 [5.3,9.2] | <0.001 |
| **WBC < 4.5, N (%)** | 83 (10.1) | 56 (7.5) | 214 (12.5) | 0.001 |
| **WBC > 11, N (%)** | 201 (24.4) | 227 (30.6) | 219 (12.8) | <0.001 |
| **IN_ICU, N (%)** | 152 (18.4) | 228 (30.7) | 39 (2.3) | <0.001 |
| **HOSPITAL, N (%)** |  |  |  |  |
| **MOUNT SINAI BI BROOKLYN** | 119 (14.4) | 200 (27.0) | 229 (13.4) | <0.001 |
| **MOUNT SINAI QUEENS HOSPITAL** | 136 (16.5) | 191 (25.8) | 197 (11.5) |  |
| **MOUNT SINAI ST. LUKE'S** | 126 (15.3) | 128 (17.3) | 302 (17.7) |  |
| **MOUNT SINAI WEST** | 78 (9.5) | 47 (6.4) | 238 (14.0) |  |
| **THE MOUNT SINAI HOSPITAL** | 366 (44.4) | 174 (23.5) | 740 (43.4) |  |
| **INFECTION STATUS, N (%)** |  |  |  |  |
| **COVID-19** | 723 (87.6) | 730 (98.4) | 1608 (94.3) | <0.001 |
| **PUI - COVID** | 102 (12.4) | 12 (1.6) | 98 (5.7) |  |
| **DAYS AFTER INFECTION, MEDIAN [Q1,Q3]** | 6.0 [2.0,11.0] | 5.0 [3.0,8.0] | 5.0 [3.0,8.0] | 0.063 |

**Effects of demographics, hospital site, and care area on time to death or discharge**. To assess the effects of individual covariates on clinical outcomes over time, we estimated the cumulative incidence functions (CIFs) for in-hospital death and discharge using a univariate competing risks survival analysis. In our full cohort, we estimated that by 10 days post-diagnosis, patients have a 51.1% chance of being recovered and being discharged from the hospital and a 21.7% chance of death (Fig S2). By grouping our cohort using different demographic and hospitalization factors, we found the differences in time-adjusted mortality among different racial groups (p=0.003, Gray’s test) is more enriched in Caucasian due to its older age group (Fig. S1B), but no differences in the CIFs of the recovered patients (p=0.408) (Fig. 2A). We also found females have better outcomes compared to males; they have trend with lower mortality rates (p=0.064) and significantly higher discharge rates (p=2.36e-4) over time (Fig. 2B), which translates to shorter in-hospital stays. Hospitalized patient outcomes also differed by hospital and care area types within hospitals, including ICUs, medical and surgical units, and other specialties (Fig. 2C-D). We found that patients admitted to hospitals in Brooklyn and Queens experienced significantly worse outcomes (Gray’s T=248.9; *p*<2e-16) compared to three hospitals based in Manhattan (Fig. 2C). This can be attributed at least partly due to the relatively older COVID-19 cohorts in Brooklyn and Queens compared with the Manhattan hospitals (Table S2). As expected, we also observed drastically worse outcomes for all patients admitted to ICU care areas compared to other care areas (Fig. 2D).

**Figure 2.**
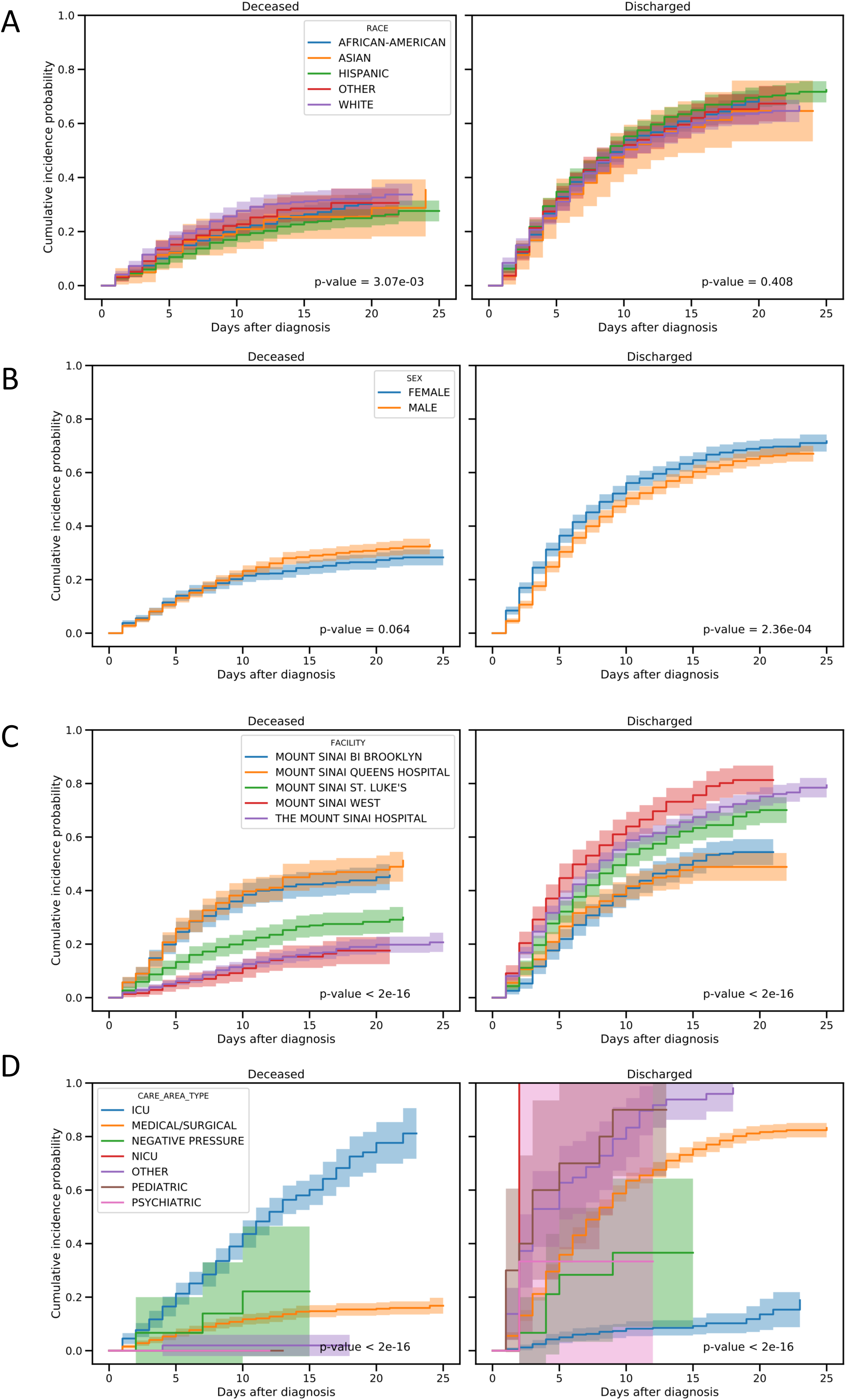
Cumulative incidence functions (CIFs) of two events, deceased and discharged, with univariate competing risks modeling. The left panels show the cumulative probability of in-hospital death for Covid-19 patients whereas the right panels show the cumulative probability of discharged after inpatient stay. The cohort was grouped by different factors including self-reported races, sex, inpatient stays at different Mount Sinai facilities and care area types, shown in rows A through D. The p-values from Gray’s test which comparing the subdistribution for deceased and discharge events across groups are shown. A significant p-values <0.05 indicates significant differences among groups in the cumulative incidence functions for the corresponding events.

**Multivariate associations between mortality and discharge**. Next, we identified etiologic factors associated with death among hospitalized patients with known outcomes (death: N = 742; discharge: N = 1,706). We found many significant associations (Fig. 3). Risk factors for death included advanced age (odds ratio, OR=1.08 [95% CI, 1.06–1.09]; p=7.07e-29), maximum temperature during hospitalization (OR=1.19 [95% CI, 1.08–1.32]; p=5.51e-4), respiratory rate >25 breaths per minute (BPM) (OR=1.74 [95% CI, 1.13–2.68]; p=0.012), ICU care area (OR=20.78 [95% CI, 12.57–35.21]; p=6.0e-31), higher WBC count (OR=1.06 [95% CI, 1.02–1.10]; p=1.02e-3), elevated serum creatinine >1.2mg/dL (OR=2.77 [95% CI, 2.05–3.75]; p=3.16e-11) and high ALT (OR=1.004 [95% CI, 1.001–1.006]; p=3.98e-3). We also found patients with higher temperature at admission (OR=0.87 [95% CI, 0.79–0.96]; p=6.65e-3), higher oxygen saturation at admission (OR=0.97 [95% CI, 0.95–1.00]; p=0.0022), as well as those with higher minimum oxygen saturation levels during hospitalization (OR=0.91 [95% CI, 0.90–0.93]; p=4.05e-25) were more likely to be discharged than be deceased (logit model OR=0.94 [95% CI, 0.91–0.97]; p=6.67e-4, Fig. 3).

**Figure 3.**
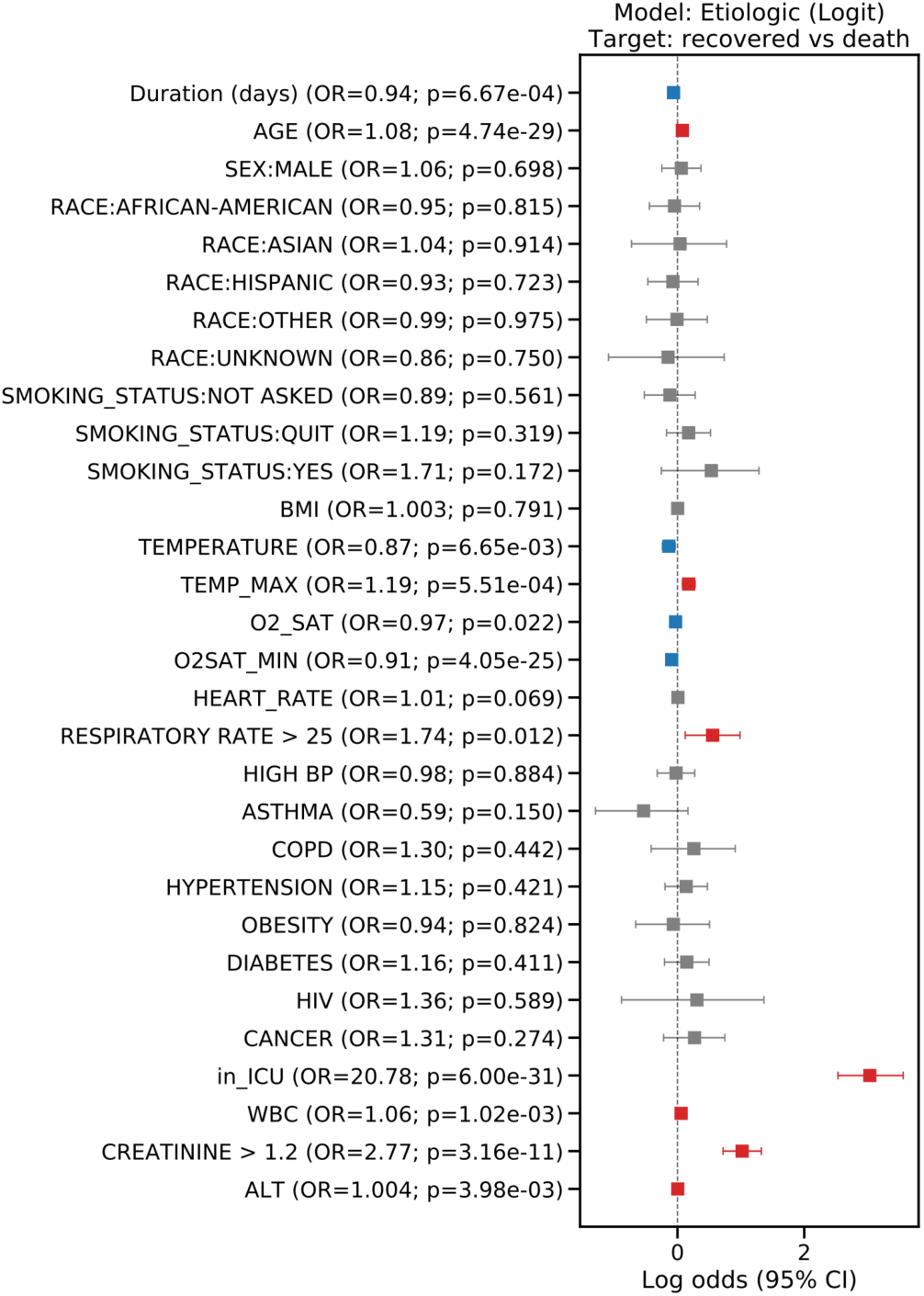
Coefficients from logistic regression models analyzing covariates associated with final outcomes (cured versus deceased) for Covid-19 patients. The estimated coefficients from the logistic regression model, also known as log odds, are plotted for the covariates. An intercept term was included in the model but excluded from the plot, which has a coefficient of −2.07 (p=0.72). Error bars indicate 95% CI.

Since *‘care in ICU area’* showed the highest OR, we then divided our samples into patients treated in the ICU or not, as these groups have very different rates of mortality (Fig. 2D). We found that only advanced age (OR=1.10 [95% CI, 1.04–1.17]; p=1.15e-3) and minimum oxygen saturation (OR=0.86 [95% CI, 0.78–0.92]; p=2.4e-4) were significantly associated with death for ICU patients (Fig. S5A), whereas all the previously discovered significant covariates persist in patients not admitted to the ICU (Fig. S5B).

**Competing risks survival analyses for inpatient mortality or discharge**. We employed a competing risks survival analysis to estimate the effects of covariates on both mortality and discharge using two Cox proportional hazard models, yielding cause-specific hazard ratios (HR) for each of the two events across all etiologic factors under consideration. This allowed us to dissect the effect of those factors on 1) reducing in-hospital mortality rate, and 2) shortening hospitalization (early discharge). Survival analyses also allowed us to leverage the additional information for patients who were still hospitalized by treating them as right-censored (i.e. the final outcome of the patients cannot be determined by the end of the study period).

The cause-specific hazards models showed advanced age significantly increased the risk of inhospital death (HR=1.05 [95% CI, 1.04-1.06]; p=1.15e-32) and decreased the probability of discharge (HR=0.98 [95% CI, 0.978-0.986]; p=4.07e-21) (Fig. 4). Minimum oxygen saturation (death: HR=0.985 [95% CI, 0.981-0.988], p=1.57e-17; discharge: HR=1.09 [95% CI, 1.08-1.10], p=2.45e-49), care in ICU (death: HR=1.58 [95% CI, 1.29-1.92], p=7.81e-6; discharge: HR=0.21 [95% CI, 0.15-0.29], p=1.06e-19), elevated creatinine (death: HR=1.75 [95% CI, 1.29-1.92], p=7.48e-10; discharge: HR=0.82 [95% CI, 0.72-0.92], p=1.22e-3) and ALT (death HR: 1.002 [95% CI, 1.001-1.003], p=8.86e-5; discharge: HR=0.998 [95% CI, 0.998-1.000], p=0.029) also all increased the risk of in-hospital death while prolonging the hospital stay (Fig. 4). Interestingly, we found that some etiologic factors only significantly influenced one outcome. For instance, BMI (death: HR=1.02, [95% CI 1.00-1.03]; p=0.021) showed a significant risk and history of COPD had a trend toward risk (death: HR=1.39, [95% CI 0.98-1.96]; p=0.066) for inhospital death, but had no significant effects on the length of hospitalization before discharge. Meanwhile, maximum temperature during hospitalization, abnormally high respiratory rate, high WBC, and history of asthma did not have significant effects on in-hospital death, but did significantly increase the length of hospital stay with HR=0.86 [95% CI, 0.82-0.89], p=5.91e-14; HR=0.66 [95% CI, 0.54-0.81], p=6.66e-5; HR=0.98 [95% CI, 0.97-1.00], p=0.011; and HR=0.78 [95% CI, 0.62-0.98], p=0.031, respectively (Fig. 4).

**Figure 4.**
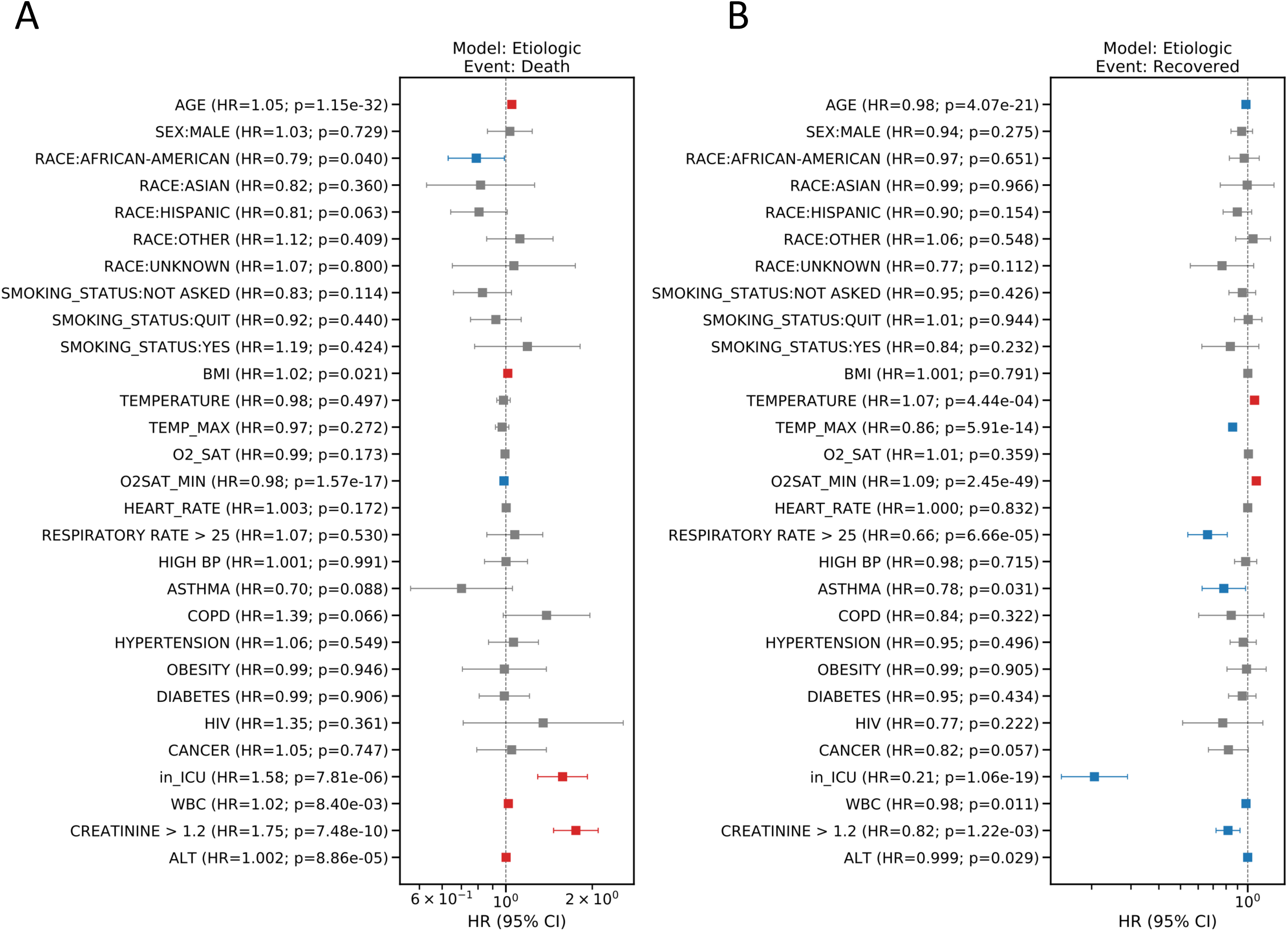
HR plots showing the results from the etiologic model. The HR from the cause-specific hazard model with competing risks (death (A) and cured (B)) are plotted for individual covariates in logarithmic scale. The estimated HR and p-values are indicated in the tick labels for those covariates. Covariates with significant elevated HR (HR > 1 and p-value<0.05) or decreased HR (HR < 1 and p-value<0.05) are highlighted in red and blue, respectively. Error bars indicate 95% CI.

**Prognostic model to forecast mortality or discharge for inpatients**. Given the large sample size, richness in clinical baseline measurements, and well-defined outcomes, we developed a prognostic model for COVID-19. Several significant predictors increased the risk for in-hospital death and decreased chance of recovery including age, BMI, oxygen saturation, elevated respiratory rate, WBC, creatinine and ALT (Fig. 5). We additionally found that Hispanic and African American patients have slightly reduced mortality rates compared to people of other races when controlling for the initial baseline physiological measurements and comorbidities (Fig. 5A). Moreover, we found history of cancer increased the length of hospitalization and delayed recovery, although it did not significantly increase risk of mortality (Fig. 5B). This prognostic model may be of significant practical value for clinicians and hospital management.

**Figure 5.**
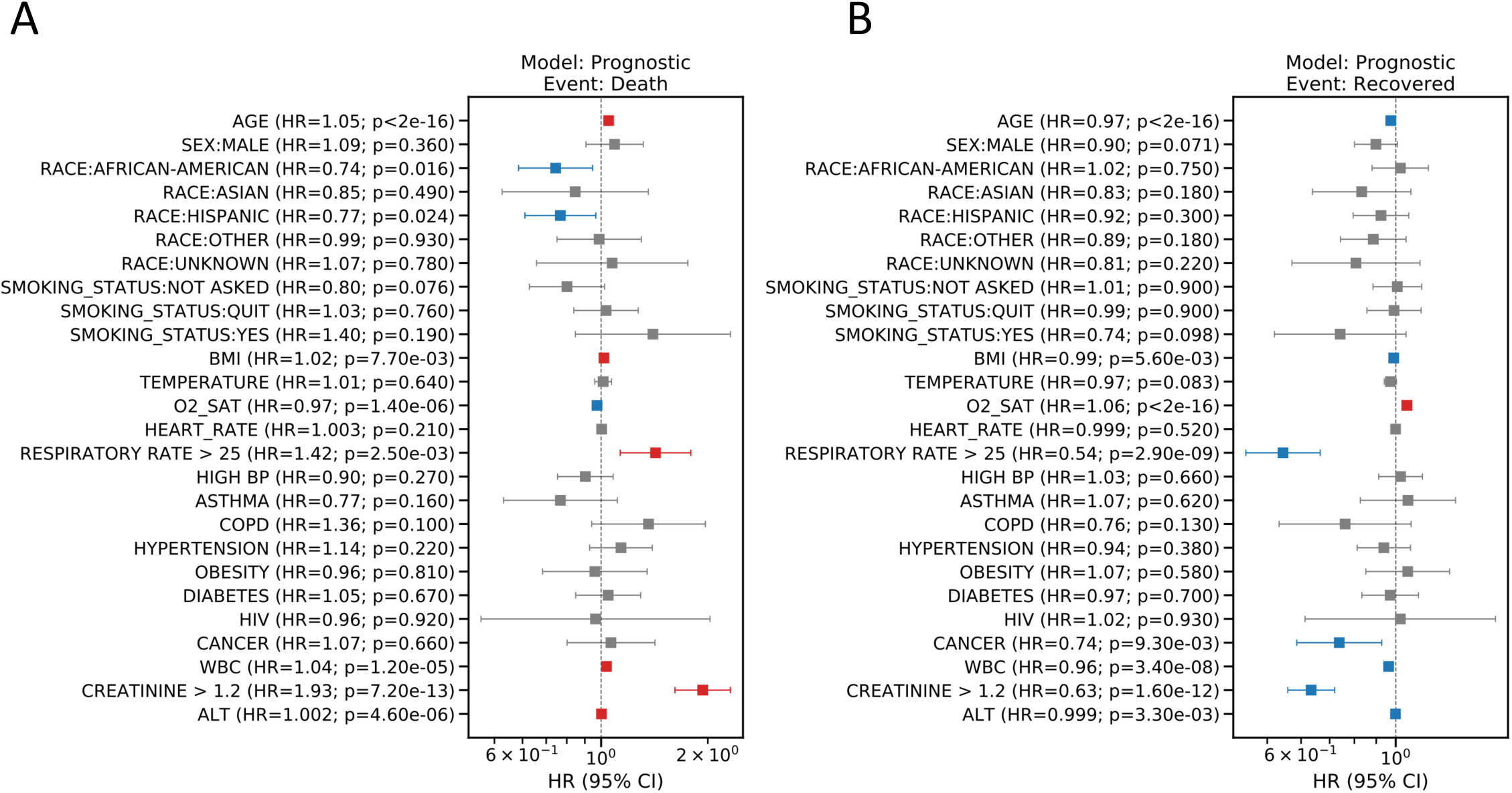
Hazard ratios (HR) plots showing the results from the prognostic model. The HR from the subdistribution hazard model with competing risks (death (A) and cured (B)) are plotted for individual covariates in logarithmic scale. The estimated HR and p-values are indicated in the tick labels for those covariates. Covariates with significant elevated HR (HR > 1 and p-value<0.05) or decreased HR (HR < 1 and p-value<0.05) are highlighted in red and blue, respectively. Error bars indicate 95% confidence intervals (CI).

## Association of elevated inflammation or biomarkers and mortality

We also assessed many laboratory tests extracted for the hospitalized COVID-19 patients in addition to WBC, creatinine and AST on their association with mortality. These laboratory tests were only available for 30% to 75% of the patients in our hospitalized cohort, therefore, are not incorporated as covariates for the previous analyses to avoid potential availability biases and decreased sample size. To estimate their associations with mortality, we fitted individual multivariate regression models for each of these lab tests controlling for age, sex and race in the corresponding subsets of cohort where these lab tests are available at baseline. Our analysis found that elevated level of LDH (OR=1.003 [95% CI, 1.002-1.004]; p=7.11e-32), CRP (OR:1.008 [95% CI, 1.006-1.01]; p=7.15e-19), D-Dimer (OR=1.16 [95% CI, 1.12-1.21]; p=1.05e-15), BUN (OR=1.02 [95% CI, 1.02-1.03]; p=1.14e-15), procalcitonin (OR=1.12 [95% CI, 1.09-1.17]; p=1.08e-10), and lower level of HGB (OR=1.07 [95% CI, 1.01-1.13]; p=0.04) are significantly associated with increased mortality (Table S3).

## Discussion

Using the largest and most racially diverse US case cohort to date, we have evaluated the impact of demographics and clinical characteristics on inpatient mortality in the Mount Sinai Health System. Among 6,158 positive or presumed positive diagnosed cases, 3,273 (50%) were admitted to one of five hospitals; of those admitted, 742 died (22%) and 1,706 recovered and discharged (52%) by the end of the study period on April 15, 2020. While we did observe higher rates of COVID-19 diagnosis among African American and Hispanic individuals, we did not observe any impact of race on mortality among inpatients. Consistent with previous reports^3,4^, we found that older individuals and men were at higher risk for mortality, as were critically ill patients cared for in the ICU. We also found that mortality varied by hospital. We identified many clinical features significantly associated with morality that may be important factors to monitor during hospital admission including respiration, temperature, heart rate, white blood cell count, creatinine, and ALT. We also estimated hazard ratios for survival, identifying oxygen saturation, ICU care, elevated creatinine and ALT as strong predictors of mortality. Finally, we developed a prognostic model to forecast risk for mortality using only baseline features, which we hope will help clinicians and hospitals identify individuals at highest risk earlier on in disease progression.

Case prevalence in African Americans and Hispanics is disproportionately high in New York^14^, which is reflected in our cohort. However, in this cohort, the disparity in positive COVID-19 diagnosis rates did not translate in our cohort to any differences in mortality, suggesting that inpatient care in the hospital does not further this disparity. While this study cannot prove it, this data suggests that there are no intrinsic biological differences which explain the racial disparities in mortality. Differences in rates of positive cases may be due to true differences in infection rates, which is consistent with higher case density in areas like Brooklyn and Queens^15^, where relatively more African American and Hispanic individuals live^16^. These areas are also more densely populated (e.g., persons per household is higher relative to Manhattan), making viral isolation more challenging^16^. It is also consistent with racial differences in occupation – African American and Hispanic residents are less likely than residents of any other race to be able to work from home, according to the Bureau of Labor Statistics^17^. This puts these groups at increased risk of infection through elevated exposure. Positive case rate differences may also in part be due to limitations on number of available tests in high case density places. According to testing rates made available by NY State (https://github.com/nychealth/coronavirus-data) combined with population estimates^15^, 3.5 residents in Brooklyn and 2.7 residents in Queens per 100 have been tested, while 4.2 per 100 have been tested in Staten Island, for example. Test availability may have resulted in biasing testing among those residents to individuals more likely to be positive, thus artificially raising the positive rate by testing fewer individuals seen as less likely to be positive (e.g., asymptomatic individuals). Fewer tests also reduce the ability to track and contain the virus, so this may contribute to higher case rates, as well. While we are encouraged to report no in-hospital differences in mortality by race, the burden of mortality will remain disproportionately held by African American and Hispanic individuals until rates of infection can be targeted and reduced.

We also found that where patients were treated affected their risk for mortality. Patients admitted to hospitals in Brooklyn or Queens were at higher risk than those at one of the Manhattan hospitals. The outer boroughs have consistently higher rates of comorbidities relevant to COVID-19 infection compared to Manhattan^18^. We found that average age in patients admitted in Brooklyn or Mount Sinai Queens was higher and oxygen saturation is lower than those in Manhattan (Table S2). Rates of comorbidities were also higher in Queens, as well (56% with any comorbidity versus 36–47% at other hospitals). Another contributing factor may be case prevalence differences by borough. There are nearly twice as many cases in Brooklyn (>31,000) and Queens (>36,000) – the counties where the Mount Sinai Brooklyn and Queens hospitals are located, respectively – than in Manhattan (>15,000)^15^. Hospitals in New York are already overwhelmed and struggling to provide sufficient staff and hospital resources to meet the need of the pandemic. It may be that the density of cases outside of Manhattan have put an additional strain on those hospitals or that only more severe patients are able to be admitted. This is an important health disparity for policymakers to address, as it may contribute to the disproportionate death of individuals living in certain neighborhoods.

The strongest predictors of hospital termination – either death or discharge – in our data were older age, higher BMI, lower oxygen saturation, care in ICU, elevated creatine and ALT levels, and history of COPD (Fig. 4). These findings were highly similar to descriptive differences reported between critically and non-critically ill hospitalized patients at NYU^3^, with the exception of COPD (not significant) and ALT (not reported). They also reported many significant differences in lab values, including C-reactive protein, d-dimer, ferritin, and procalcitonin. These lab tests were not routinely collected on all of our patients, however, among patients with available measures, we were able to corroborate their evidence that higher baseline measures of all aforementioned labs were associated with mortality (Table S3). Given the consistency of these findings, we suggest oxygen saturation, creatine, C-reactive protein, d-dimer, ferritin, and procalcitonin are good targets to monitor throughout hospital admission as they are sensitive to clinical outcome across reported data. We additionally found that high temperature, abnormally high respiratory rate, high WBC, and history of asthma significantly lengthened hospital stays (Fig. 4), suggesting that these vital signs and labs may also be of use for monitoring disease progression and severity.

One major reason for high mortality rates from COVID-19 is that no proven effective therapies exist as yet^6^. Many new treatments have been quickly developed or adapted, and despite incomplete evidence of efficacy, have been incorporated into current clinical practice. For instance, heparin was administered as part of standard of care at Mount Sinai as of April 10, 2020 due to the negative impact of SARS-CoV-2 on coagulation and consequent professional recommendations^19^. Therefore, the efficacy of the use of heparin is limited based upon the data availability and subsequent follow up. While results from ongoing clinical trials are still unknown, there is an immediate need for any useful information on patient response to these pharmacologic treatments.

Finally, we discovered several significant baseline predictors increasing risk for in-hospital death: older age, higher BMI, lower oxygen saturation, elevated respiratory rate, elevated WBC, elevated creatinine and elevated ALT (Fig. 5). Assessing these patient characteristics immediately upon hospital admission may help identify individuals at the highest risk and help determine clinical action. We hope this prognostic model may be of significant practical value for clinicians and hospital management.

Our study should be considered in light of many limitations. First, many patients were missing key inflammatory markers like C-reactive protein and d-dimer, so we could not include them in our multivariate models without losing significant power. We have provided results for these labs analyzed individually (Table S4) that are in line with other reports^3^, however future work incorporating these labs with other measures would yield a more comprehensive ranking of significant indicators of mortality. Additionally, clinical course trajectories of lab values and vital signs across hospitalization are likely to give valuable insight into disease severity and progression, but analysis at this level requires substantially more repeated measurements per patient than were available at the time of this study. We also expect outcomes to change as local outbreaks become contained (or not) and additional information on novel treatments is made available. However, given the urgent need for prognostic indictors amidst the ongoing pandemic, we believe this report, the largest and more diverse population to date, provides an important initial summary of clinical features associated with mortality and can facilitate risk assessment and care. While further studies are imperative, our ability to use modeling techniques to assimilate and analyze large data sets quickly and efficiently provide a complimentary approach as we await vital data from clinical trials. We anticipate that emerging reports across the country will be combined and compared, as health-related behaviors (both personal and mandated by state and local governments), as well as hospital protocols and resources vary considerably.

In this study, we estimated the effect of key clinical characteristics on mortality among patients hospitalized in one of five Mount Sinai hospitals in New York, Brooklyn, and Queens. Based on these findings, first, we recommend considering for hospital admission patients with the following characteristics: older age, higher BMI, lower oxygen saturation, elevated respiratory rate, and elevated lab parameters (WBC, creatinine and ALT) as prognostic indicators for increased risk for mortality. Second, we identified changes in respiration, temperature, heart rate, white blood cell count, creatinine, and ALT as particularly important features to monitor during hospital admission to track risk for increased mortality. Together, we hope these estimates can help inform clinicians and hospitals early on which patients are at greatest risk, what ongoing clinical features track with disease progression.

## Data Availability

Individual level data are not available for this study. For aggregate data please contact the corresponding authors.

## Acknowledgements

We would like to express our sincerest condolences to the patients and their families who underwent from the COVID-19 outbreak. We greatly appreciate the all medical staff who worked together to overcome the COVID-19 outbreak. We are grateful of research opportunity from Dr. Eric Nestler. We also thank the helpful comments from Dr. Emma Benn.

## Author Contributions

Li, Wang had full access to all of the data in the study and takes responsibility for the integrity of the data and the accuracy of the data analysis.

## Concept and design

Li, Chen

## Acquisition, analysis, or interpretation of data

Wang, Kao, Li, Chen, Oh, Schadt, Kovatch, Nirenberg, Zheutlin, Gross

## Drafting of the manuscript

Wang, Zheutlin, Li, Oh, Gross, Chen

## Critical revision of the manuscript for important intellectual content

Wang, Zheutlin, Li, Chen, Oh, Schadt, Gross, Ayers, Glicksberg, A. Charney, Nadkarni, O’Reilly, Just, Horowitz, D. Charney, Reich

## Statistical analysis

Wang, Kao, Ayers, Li, Chen

## Administrative, technical, or material support

Li, Wang, Kao, Zheutlin, Kovatch, Nirenberg, Martin, Schadt

## Supervision

Li, Chen, Schadt

---

**Table S1.** Estimated coefficients from age-adjusted logistic regression model showing the racial disparities in the infection rates.

|  | **ESTIMATED**<br>**COEFFICIENT** | **STD**<br>**ERR** | **Z** | **P>\|Z\|** | **2.5%**<br>**CI** | **97.5%**<br>**CI** |
| --- | --- | --- | --- | --- | --- | --- |
| **(INTERCEPT)** | -7.8584 | 0.046 | -172.362 | 0 | -7.948 | -7.769 |
| **AGE** | 0.0345 | 0.001 | 53.074 | 0 | 0.033 | 0.036 |
| **AFRICAN-AMERICAN/BLACK** | 0.643 | 0.034 | 18.87 | 0 | 0.576 | 0.71 |
| **ASIAN/PACIFIC-ISLANDER** | 0.0159 | 0.06 | 0.263 | 0.793 | -0.103 | 0.134 |
| **HISPANIC/LATINO** | 1.9757 | 0.033 | 60.152 | 0 | 1.911 | 2.04 |
| **OTHER** | 0.6316 | 0.04 | 15.911 | 0 | 0.554 | 0.709 |

**Table S2.** Descriptive statistics of the age and baseline O2 saturation at admission of hospitalized COVID-19 patients by hospitals.

| **FACILITY** | **AGE MEAN** | **AGE STD** | **O2 SAT**<br>**BASELINE**<br>**MEAN** | **O2 SAT**<br>**BASELINE**<br>**STD** |
| --- | --- | --- | --- | --- |
| **MOUNT SINAI BI BROOKLYN** | 69.73 | 13.82 | 94.41 | 7.78 |
| **MOUNT SINAI QUEENS**<br>**HOSPITAL** | 65.16 | 15.39 | 91.26 | 8.89 |
| **MOUNT SINAI ST. LUKE'S** | 66.58 | 16.41 | 93.57 | 6.94 |
| **MOUNT SINAI WEST** | 62.07 | 18.68 | 93.59 | 5.87 |
| **THE MOUNT SINAI**<br>**HOSPITAL** | 58.88 | 17.77 | 94.20 | 5.80 |

**Table S3.** p-values and odd ratios (OR) for less common baseline laboratory tests in multivariate association analyses between COVID-19 mortality and discharge.

| **LABORATORY TESTS** | **P-VALUE** | **LOG**<br>**ODDS** | **ODDS**<br>**RATIO** | **2.5%**<br>**OR** | **97.5%**<br>**OR** |
| --- | --- | --- | --- | --- | --- |
| **LDH** | 7.11E-32 | 0.0030 | 1.0030 | 1.0025 | 1.0035 |
| **CRP** | 7.15E-19 | 0.0080 | 1.0080 | 1.0063 | 1.0098 |
| **D-DIMER** | 1.05E-15 | 0.1524 | 1.1646 | 1.1228 | 1.2097 |
| **BUN** | 1.14E-15 | 0.0235 | 1.0238 | 1.0181 | 1.0299 |
| **PROCALCITONIN** | 1.08E-10 | 0.1153 | 1.1222 | 1.0858 | 1.1652 |
| **FERRITIN** | 2.07E-09 | 0.0001 | 1.0001 | 1.0001 | 1.0002 |
| **FIBRINOGEN** | 2.89E-03 | 0.0012 | 1.0012 | 1.0004 | 1.0020 |
| **HGB** | 4.08E-02 | -0.0683 | 0.9340 | 0.8748 | 0.9973 |
| **EGFR NON-AFRICAN AM** | 1.07E-01 | -0.0080 | 0.9920 | 0.9823 | 1.0017 |
| **EGFR AFRICAN AM** | 5.21E-01 | 0.0035 | 1.0035 | 0.9927 | 1.0145 |

**Figure S1.**
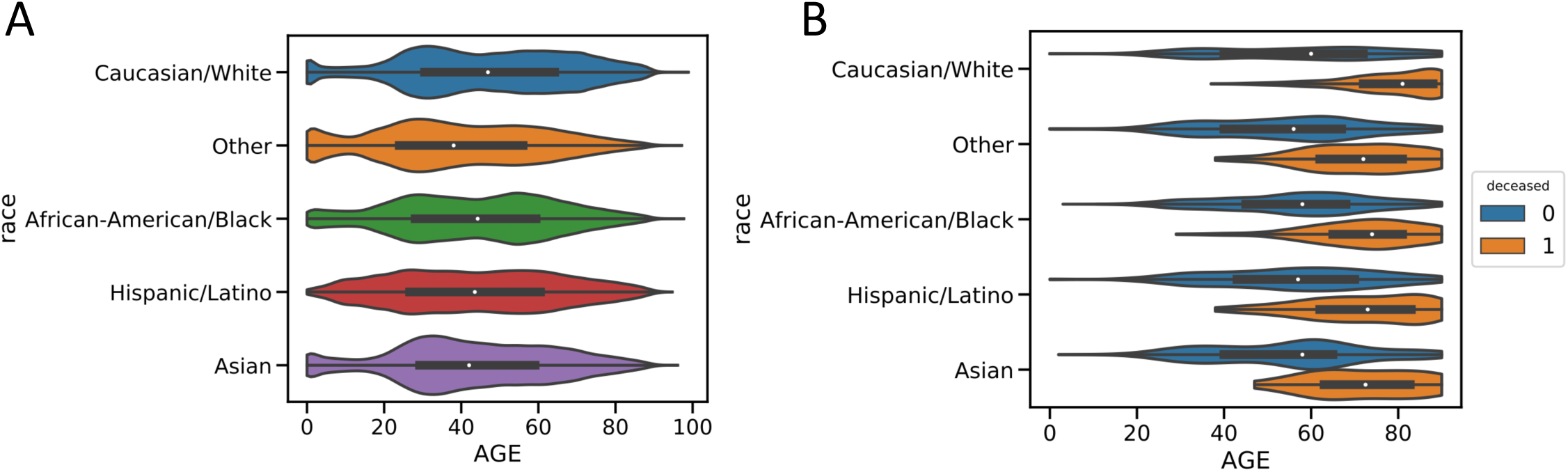
Age distributions across racial groups of patients from the MSHS EMR (A) and Covid-19 patients (B).

**Figure S2.**
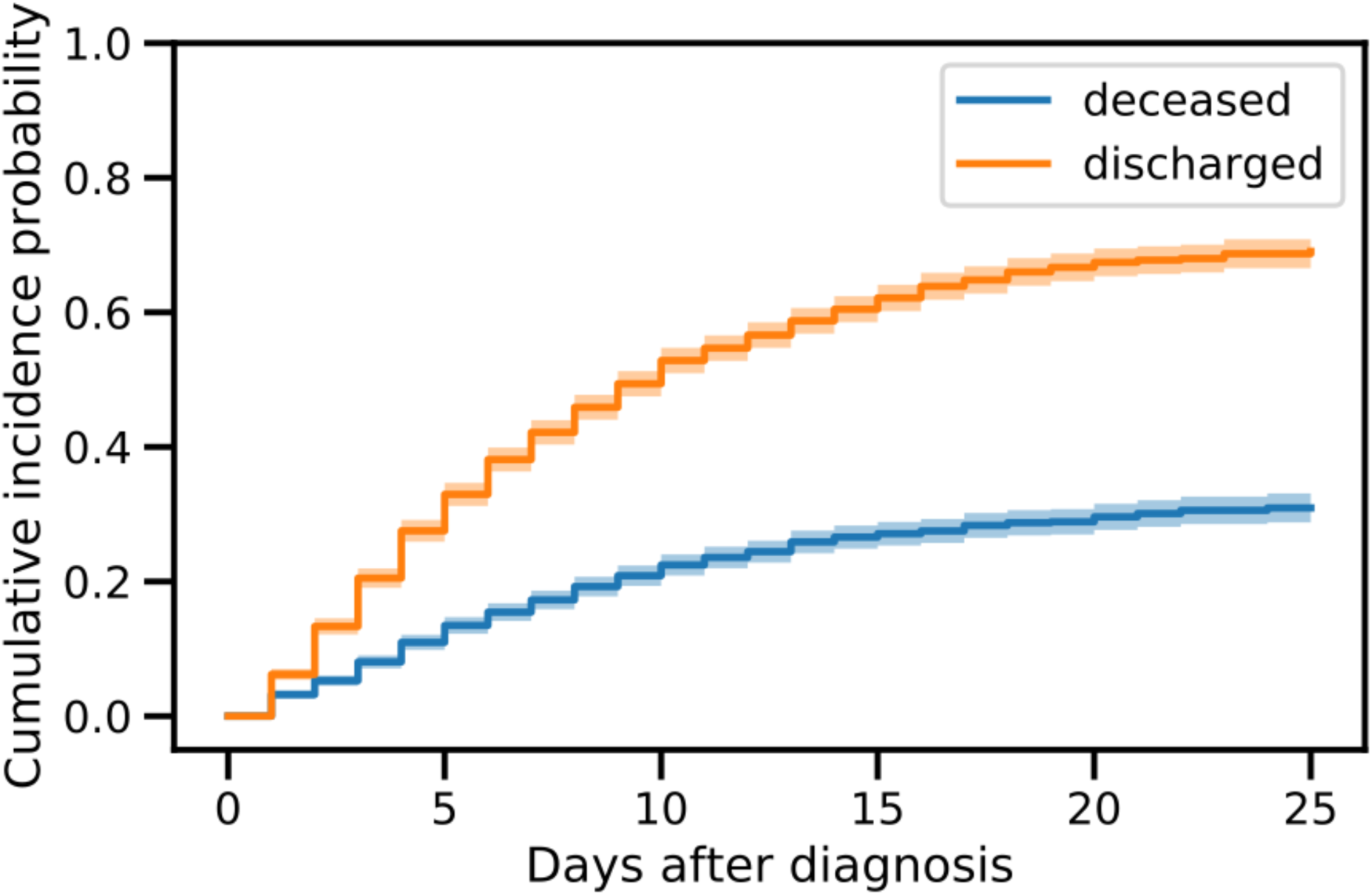
Overall CIFs for the hospitalized Covid-19 cohorts for deceased and discharged events, respectively.

**Figure S3.**
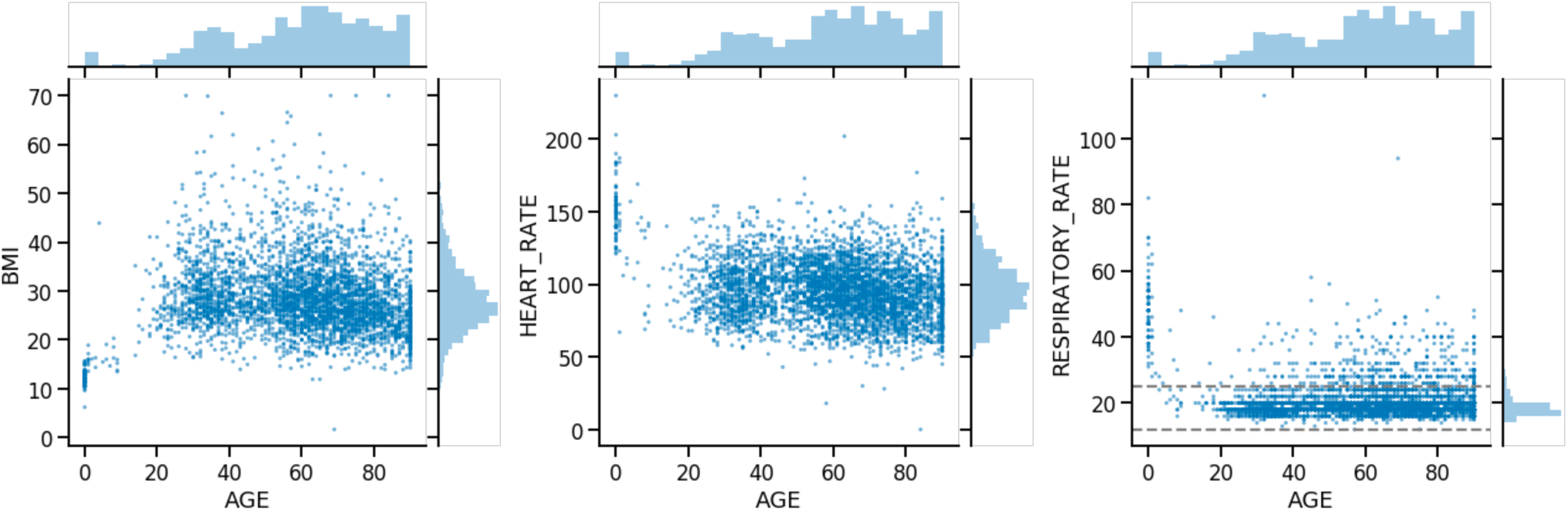
Scatter plots showing various vital signs with respect to age of the hospitalized Covid-19 cohort.

**Figure S4.**
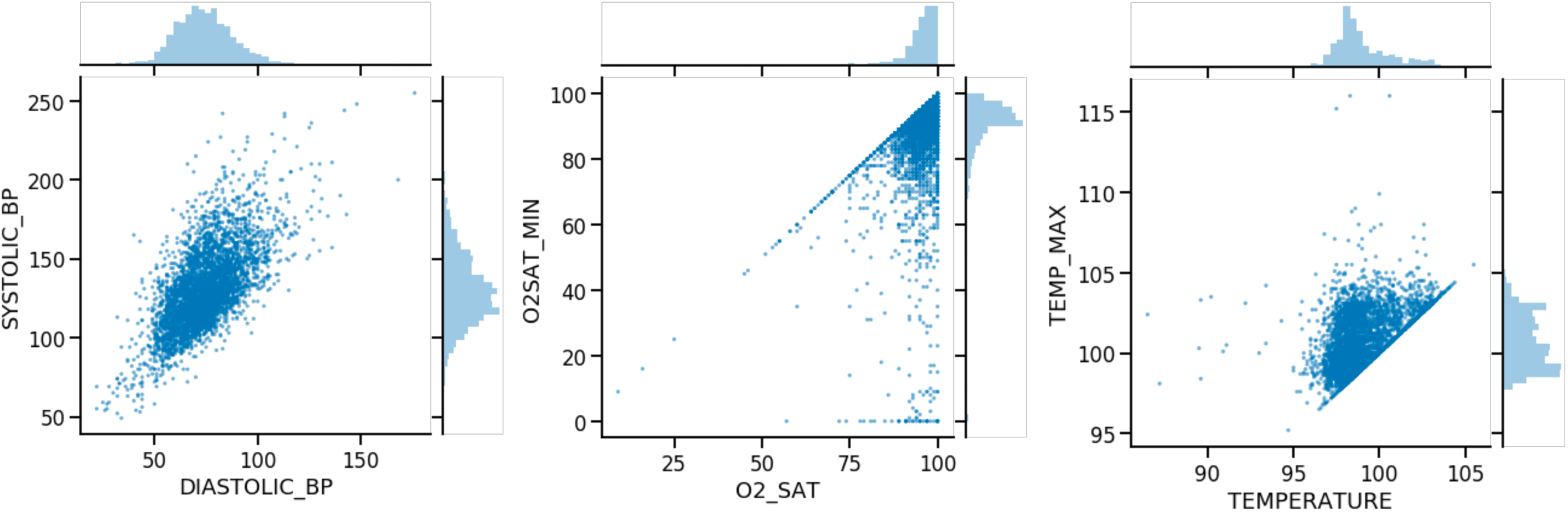
Scatter plots showing related vital signs against each other.

**Figure S5.**
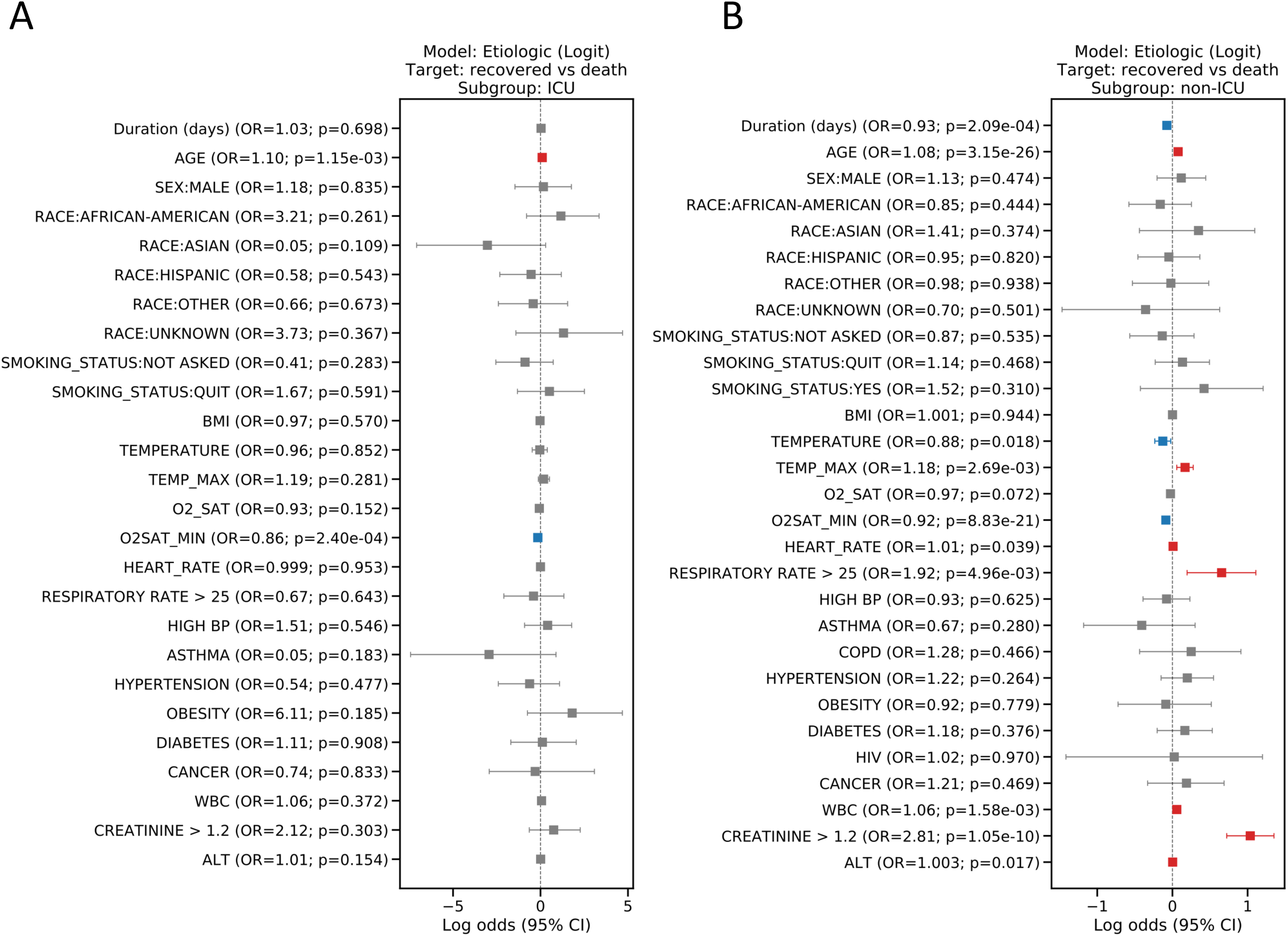
Coefficients from logistic regression models analyzing covariates associated with final outcomes (cured versus deceased) for COVID-19 patients with and without ICU stays (A, B). The estimated coefficients from the logistic regression model, also known as log odds, are plotted for the covariates. An intercept term was included in the model but excluded from the plot. Error bars indicate 95% CI.

**Figure S6.**
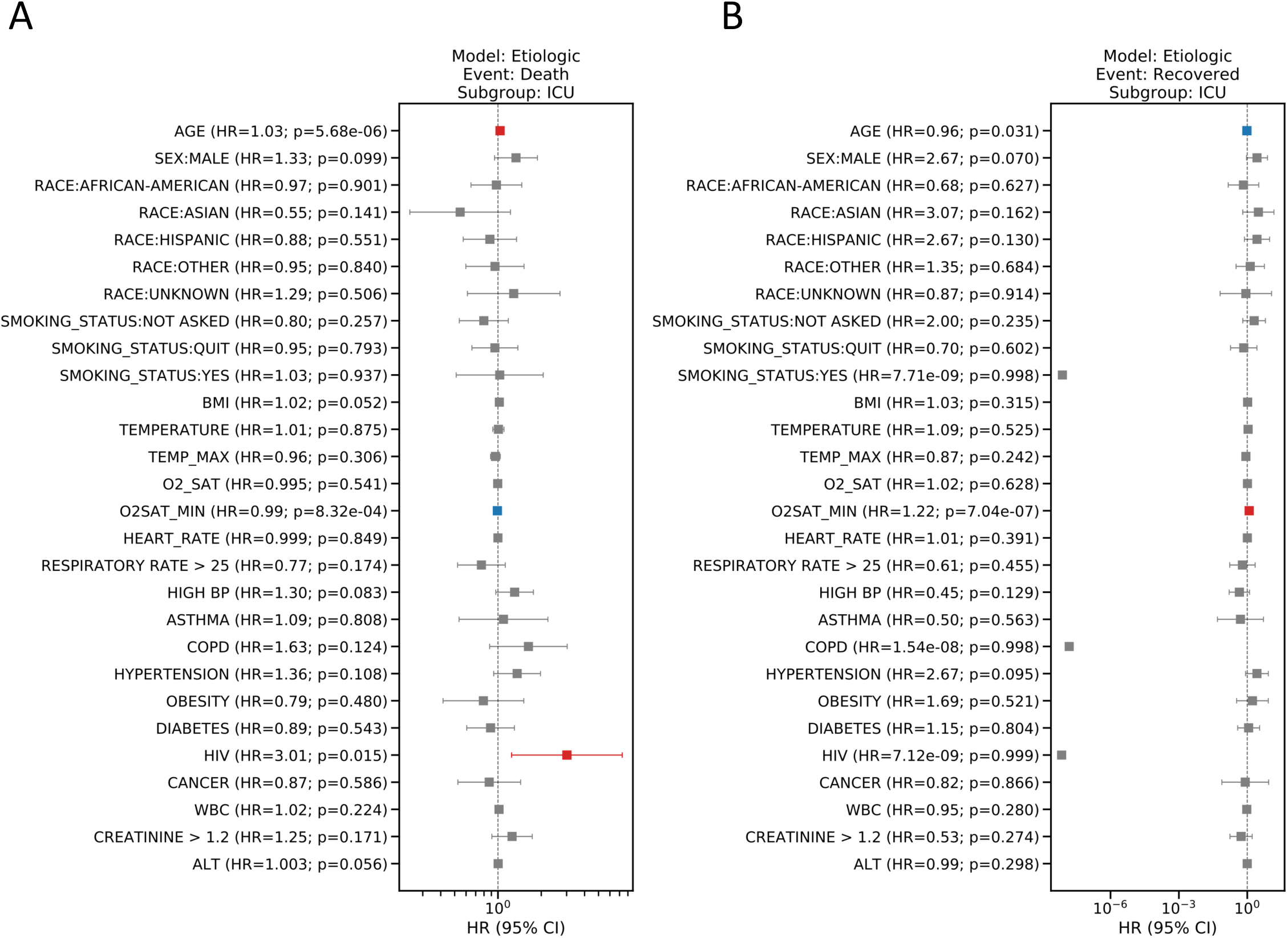
HR plots showing the results from the etiologic model exclusively for Covid-19 patients with ICU stays. The HR from the cause-specific hazard model with competing risks (death (A) and cured (B)) are plotted for individual covariates in logarithmic scale. The estimated HR and p-values are indicated in the tick labels for those covariates. Covariates with significant elevated HR (HR > 1 and p-value<0.05) or decreased HR (HR < 1 and p-value<0.05) are highlighted in red and blue, respectively. Error bars indicate 95% CI.

**Figure S7.**
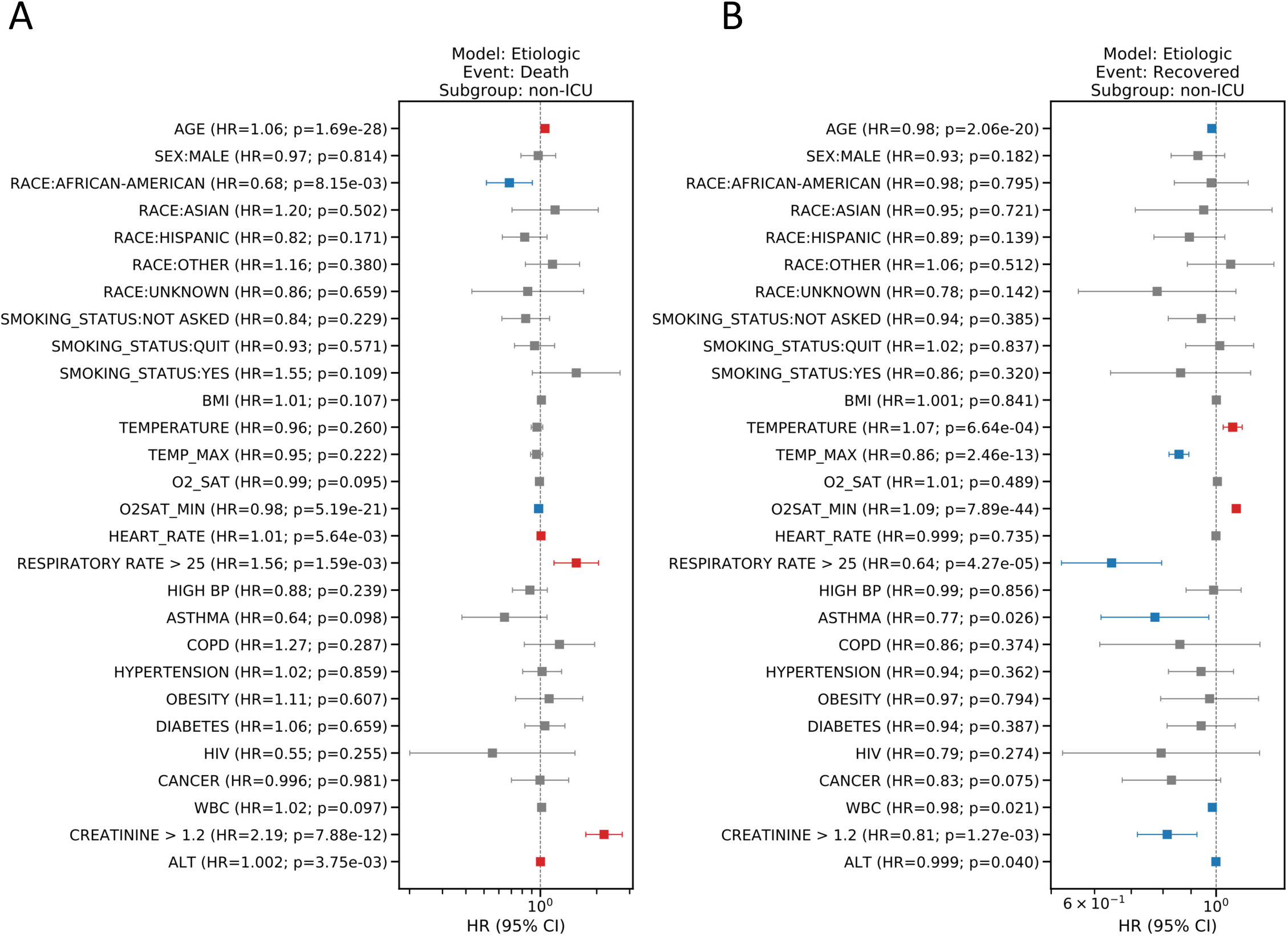
HR plots showing the results from the etiologic model exclusively for Covid-19 patients without ICU stays. The HR from the cause-specific hazard model with competing risks (death (A) and cured (B)) are plotted for individual covariates in logarithmic scale. The estimated HR and p-values are indicated in the tick labels for those covariates. Covariates with significant elevated HR (HR > 1 and p-value<0.05) or decreased HR (HR < 1 and p-value<0.05) are highlighted in red and blue, respectively. Error bars indicate 95% CI.

## References

1. Gudbjartsson DF, Helgason A, Jonsson H, et al. Spread of SARS-CoV-2 in the Icelandic Population. N Engl J Med. 2020.

2. Tang W, Cao Z, Han M, et al. Hydroxychloroquine in patients with COVID-19: an open-label, randomized, controlled trial. *medRxiv*. 2020:2020.2004.2010.20060558.

3. Petrilli CM, Jones SA, Yang J, et al. Factors associated with hospitalization and critical illness among 4,103 patients with COVID-19 disease in New York City. *medRxiv*. 2020.

4. Garg S. Hospitalization Rates and Characteristics of Patients Hospitalized with Laboratory-Confirmed Coronavirus Disease 2019—COVID-NET, 14 States, March 1–30, 2020. MMWR Morbidity and Mortality Weekly Report. 2020;69.

5. Yancy CW. COVID-19 and African Americans. JAMA. 2020.

6. Sanders JM, Monogue ML, Jodlowski TZ, Cutrell JB. Pharmacologic Treatments for Coronavirus Disease 2019 (COVID-19): A Review. JAMA. 2020.

7. Austin PC, Lee DS, Fine JP. Introduction to the analysis of survival data in the presence of competing risks. Circulation. 2016;133(6):601–609.

8. Wolbers M, Koller MT, Stel VS, et al. Competing risks analyses: objectives and approaches. European heart journal. 2014;35(42):2936–2941.

9. Brock GN, Barnes C, Ramirez JA, Myers J. How to handle mortality when investigating length of hospital stay and time to clinical stability. BMC medical research methodology. 2011;11(1):144.

10. Gray RJ. A class of K-sample tests for comparing the cumulative incidence of a competing risk. The Annals of statistics. 1988: 1141–1154.

11. Fine JP, Gray RJ. A proportional hazards model for the subdistribution of a competing risk. Journal of the American statistical association. 1999;94(446):496–509.

12. Cox DR, Oakes D. Analysis of survival data. Vol 21: CRC Press; 1984.

13. Gray B, Gray MB, Gray R. The cmprsk package. The Comprehensive R Archive network. 2004.

14. Health NDo. NYC Health COVID-19: Data. https://www1.nyc.gov/site/doh/covid/covid-19-data.page. Published 2020. Accessed 04/15/2020.

15. Dong E, Du H, Gardner L. An interactive web-based dashboard to track COVID-19 in real time. The Lancet infectious diseases. 2020.

16. Bureau UC. Census quick facts. https://www.census.gov/quickfacts/fact/table/newyorkcitynewyork,bronxcountybronxboroughnewyork,kingscountybrooklynboroughnewyork,newyorkcountymanhattanboroughnewyork,queenscountyqueensboroughnewyork,richmondcountystatenislandboroughnewyork/PST045218. Published 2020. Accessed 04/15/2020.

17. Labor UDo. Job Flexibilities And Work Schedules — 2017–2018. 09/24/2019 2019.

18. Health N. NYC Data: Community Health Profiles. https://a816-health.nyc.gov/hdi/profiles/. Accessed 04/19/2020.

19. Thachil J, Tang N, Gando S, et al. ISTH interim guidance on recognition and management of coagulopathy in COVID-19. Journal of Thrombosis and Haemostasis.n/a(n/a).

